# Efficacy of Ivermectin in COVID-19 Patients with Mild to Moderate Disease

**DOI:** 10.1101/2021.02.02.21250840

**Authors:** Karamat Hussain Shah Bukhari, Asma Asghar, Najma Perveen, Arshad Hayat, Sermad Ahmad Mangat, Kamil Rehman Butt, Mohammad Abdullah, Tehreem Fatima, Ahmad Mustafa, Talal Iqbal

## Abstract

**Objective:** To evaluate the efficacy of ivermectin (IVM) as an addition to the standard of care (SOC) treatment in COVID-19 patients with mild and moderate disease

**Materials and Methods:** A randomized clinical trial (Trial registration # NCT04392713) was carried out at Combined Military Hospital Lahore from March 15, 2020, to June 15, 2020. Eighty-six patients with reverse transcriptase-polymerase chain reaction (RT-PCR) proven SARS-CoV-2 infection completed the trial protocol. Patients were stratified via the lottery method into two groups. Group A was administered standard of care (SOC) treatment as per existing hospital guidelines whereas group B was given ivermectin (single dose of 12 milligrams) along with SOC treatment. PCR was repeated at 72 hours, 7^th^ day, and at 14^th^ day of admission for both the groups and the point at which the PCR became negative was noted. Complete blood counts, liver function tests and renal function tests were done at recruitment, 7^th^ day, and 14^th^ day. The primary outcome was the viral clearance, measured as days to achieve PCR negativity. The secondary outcome was the development of any adverse side effects pertinent to ivermectin or derangement in baseline laboratory parameters.

**Results:** In group A, 36 (80%) participants were males, and 9 (20%) were females, whereas in group B, 37 (90.2%) were males and 4 (9.8%) were females. Mean age was 39.0± 12.6 and 42.2 ± 12.0 years for groups A and B, respectively (p= 0.394). There was early viral clearance in group B as compared to group A (p=0.001). No adverse reaction or derangements in laboratory parameters was noted in the intervention arm during the trial period.

**Conclusion:** In the intervention arm, early viral clearance was observed and no side effects were documented. Therefore ivermectin is a potential addition to the standard care of treatment in COVID-19 patients.

## Introduction

The COVID-19 pandemic has not only impacted the health of the masses but also has had vast economic, psychological, and social implications. Due to the extensive burden of disease on healthcare facilities and healthcare providers, it is the need of the hour to find a therapy that is risk-free and economically viable. Globally, numerous trials are underway to find therapeutic options.

Ivermectin belongs to the compound group called avermectin, named after their microorganism origin, Streptomyces avermitilis [1]. It has primarily been used as an anti-helminthic drug owing to its action on the gamma-aminobutyric acid (GABA) gated chloride channels that cause muscle hyperpolarization of muscle and neurons, eventually leading to death in parasites [2]. It also has anti-viral properties due to its action on importin α/β1 mediated nuclear transport. Ivermectin prevents the binding of viral proteins to importin α/β1 rendering the viral proteins unable to enter the nucleus and cause infection [3]. After ingestion, ivermectin is rapidly absorbed and reaches maximum blood levels in four hours. It also has a high affinity for plasma proteins and has a large volume of distribution (≈ 3.5liters/kg), hence the drug stays in the body for a longer time, which translates as a persistent drug effect [4].

The anti-viral effects of ivermectin have been documented against several RNA viruses such as Zika, dengue, yellow fever, chikungunya, and HIV-1. [5]. In a recent in vitro study, Caly et al. demonstrated a 5000-fold reduction in the SARS-CoV-2 viral RNA within 48 hours of ivermectin administration [6]. In COVID-19, a high viral load is invariably linked to the disease taking a complicated course and may lead to cytokine release syndrome (CRS). High virus titer and the subsequent strong inflammatory cytokine response are related to high morbidity and mortality. The experience from treating severe acute respiratory syndrome (SARS) and the Middle East respiratory syndrome (MERS) shows that reducing viral load early on may improve disease prognosis [7, 8].

All these aforementioned factors make it a front runner in the treatment of COVID-19 and the drug is already being studied in over 70 clinical trials all over the world [9]. The objective of this trial is to establish the efficacy of ivermectin for COVID-19 patients with mild to moderate disease.

## Materials & Methods

This study is a randomized controlled clinical trial (Trial Registration # NCT04392713) carried out from 15th March to 15th June 2020 after approval by the institutional review board and was conducted in compliance with the principles laid down in the Declaration of Helsinki.

The trial enrolled patients aged 15-65 years of both genders who were COVID-19 positive, proven by reverse transcriptase-polymerase chain reaction (RT-PCR) and having mild to moderate severity of the disease. Patients were considered eligible if they were able to consent for trial, stated their willingness to comply with all study procedures, agreed for admission for the trial period (14 days), and those able to take oral medication. All females of childbearing age were to undergo a pregnancy test and a positive test would mean that they would be excluded. Patients with severe symptoms, likely due to cytokine release syndrome, those with uncontrolled co-morbidities, and immunocompromised states were excluded. Patients with a history of ivermectin allergy were also excluded. Drug history was also sought, and patients taking CYP 3A4 inhibitors or inducers were also excluded. A chest x-ray (CXR) was used to support “moderate” severity and if these patients had oxygen requirements equivalent to FiO2 ≥ 50%, they were excluded. The severity of the disease was defined by the World Health Organization (WHO) guidelines (Table *1*). WHO classifies the disease into mild, moderate, severe, and critical disease.

**Table 1:** World Health Organization Severity Score for COVID-19 Patients [10].

| Types | Findings |
| --- | --- |
| **Mild** | Mild clinical symptoms [fever < 38°C (quelled without treatment) with or without cough, no dyspnea, no gasping, no chronic disease)<br>No imaging findings of pneumonia. |
| **Moderate** | Fever, respiratory symptoms, imaging findings of pneumonia |
| **Severe** | Meet any of the followings:<br>• Respiratory distress RR≥30 times/min<br>• SpO<sub>2</sub> < 93% at rest<br>• PaO<sub>2</sub>/FiO<sub>2</sub> ≤ 300 mmHg<br>Patients showing a rapid progression (>50%) on CT imaging within 24-48 hours should be managed as severe (added in the trial sixth edition) |
| **Critical** | Meet any oh the following:<br>• Respiratory failure, need medical assistance<br>• Shock<br>• “Extrapulmonary” organ failure, intensive care unit is needed. |

The primary end-point of the study was viral clearance and was measured as the days to achieve RT-PCR negativity following ivermectin administration. The secondary outcome was the development of any adverse side effects pertinent to ivermectin or derangement in baseline laboratory parameters.

The enrolled patients were allocated to two groups; either to group A (the control arm) or to group B (the intervention arm) as shown in Figure *1*. The patients were randomized in a 1:1 ratio via a lottery method. Group A was to receive standard of care (SOC) alone. The SOC included oral vitamin C 500mg once daily, oral vitamin D3 200,000 IU once weekly, and oral paracetamol 500 mg SOS. Group B received SOC and a single dose of ivermectin 12 milligrams (mg) at admission. There was no blinding, and patients in the intervention arm were provided with information about the drug, and their informed consent was taken.

**Figure 1:**
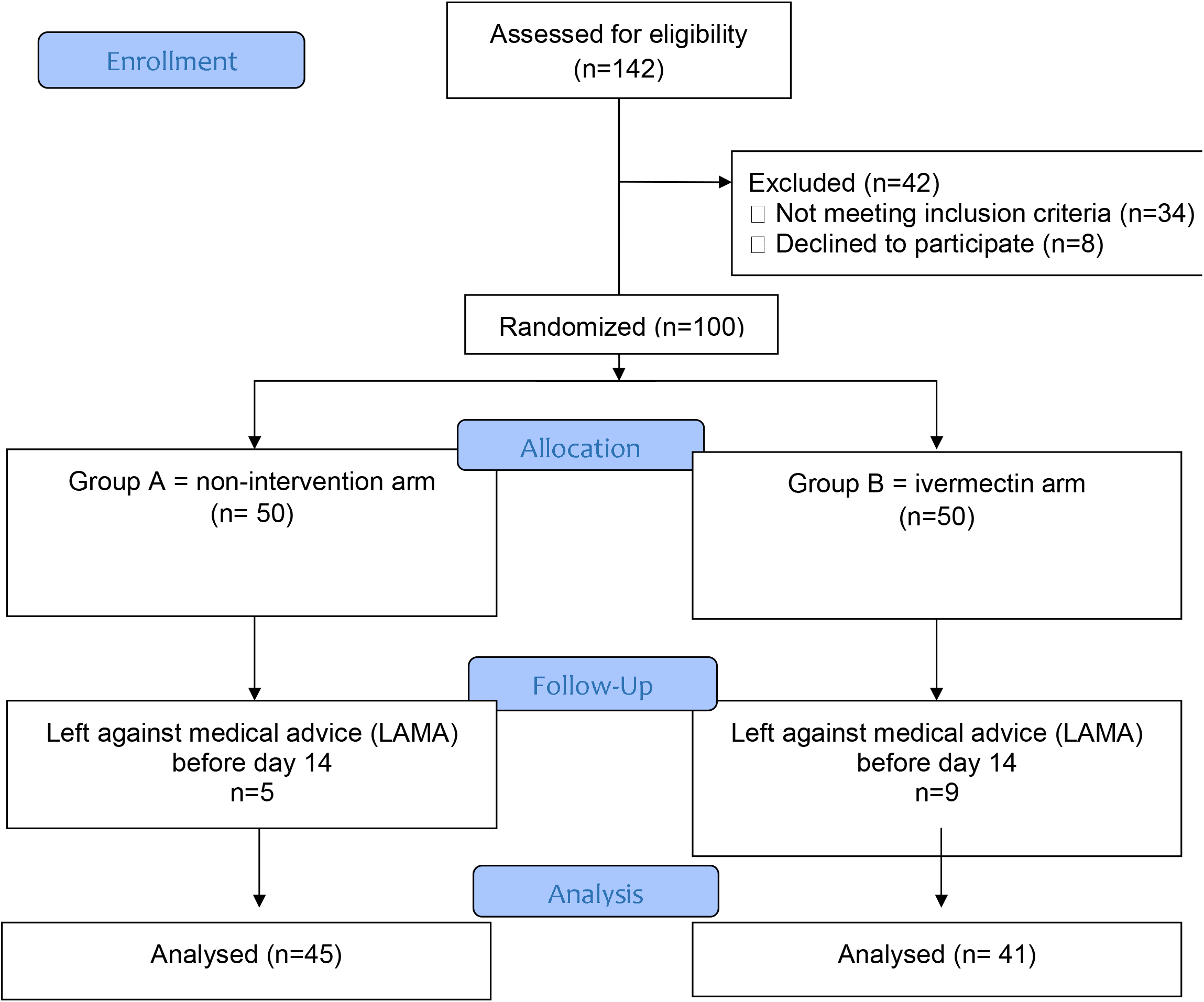
CONSORT diagram for the trial.

SARS-CoV-2 RT-PCR was repeated at 72 hours, on day 7, and day 14 post-admission. Complete blood count, renal and liver function tests were done at recruitment day, 72 hours, at day 7, and at day 14, to monitor for derangements in any lab parameters. Adverse reactions (i.e., pruritus, fever, rash, myalgia, headache), ocular symptoms, gastrointestinal symptoms, neurological symptoms, joint problems were monitored daily. Patients were discharged after 14 days with a single negative PCR result or when they had 2 negative PCR results. All patients were then followed up with a telephonic interview on the 28th day and questions pertaining to general health and the possible development of any side effects to ivermectin were inquired.

Demographic and baseline characteristics, including age, gender, and co-morbidities were noted for all patients. The data was entered and analyzed using SPSSv25 for relevant statistical tests of significance at a 95% confidence interval and p-values ≤ 0.05 were considered significant.

## Results

The trial commenced with a total of 100 patients, from which 14 patients (5 from group A and 9 from group B) dropped out and/or left against medical advice before the study period could have been completed. Out of the final total of 86 participants, group A (control arm) comprised of 45 patients, while group B (intervention arm) had 41. Clinical and demographic features have been shown in Table *2*.

**Table 2:** Demographics and clinical features of the study population.

|  | **Group A**<br>**(n=45)** | **Group B**<br>**(n=41)** | **P-value** |
| --- | --- | --- | --- |
| Mean age ± SD (years) | 38.98 ± 12.61 | 42.24 ± 12.0 | 0.394 |
| Male | 36 (80%) | 37 (90.2%) | 0.185 |
| Female | 9 (20%) | 4 (9.8%) |  |
| Diabetes mellitus | 4 (8.9%) | 6 (14.6%) | 0.406 |
| Hypertension | 7 (15.6%) | 5 (12.2%) | 0.653 |
| Ischemic heart disease | 2 (4.4%) | 3 (7.3%) | 0.57 |

RT-PCR was repeated at 72 hours, on day 7, and day 14 post-admission and the time to PCR negativity was noted. There was early viral clearance in the intervention arm in terms of PCR negativity (Table *3*).

**Table 3:** Patients with negative PCR at various stages of the trial.

|  | **72 hours** | **Day 7** | **Day 14** | **P-value** |
| --- | --- | --- | --- | --- |
| Group A<br>(control arm) | 2 | 18 | 25 | 0.001 |
| Group B<br>(intervention arm) | 17 | 20 | 4 |  |

No untoward drug reactions were recorded in any patient throughout the trial period (14 days) or on telephonic follow-up on the 28th day. We inquired about the development of any pruritus, fever, rash, myalgia, headache, ocular symptoms, gastrointestinal symptoms, neurological symptoms, and joint problems.

## Discussion

Clinically, COVID-19 can present from an asymptomatic carrier state to life-threatening pulmonary involvement requiring mechanical ventilation. In diseases with a broad clinical spectrum, to better understand the effectiveness of any intervention it’s important to study a sub-population of patients, and this is why we only choose patients with mild to moderate disease. In this randomized clinical trial, we noted that the use of ivermectin is linked to early viral clearance in patients with mild to moderate COVID-19 disease. Since the recent success by Caly et al. [6] to reduce SARS-CoV-2 viral load by ivermectin in vitro, there has been a great stir and hunt to validate this in vivo.

In response to Caly et al., Craig et al [11] made a physiologic-based pharmacokinetic (PBPK) model using the Simcyp platform to explore plasma and lung concentrations of ivermectin and they were successful in achieving IC50. Furthermore, the retrospective ICON (Ivermectin in COvid-19 Nineteen) study (n=283) showed that patients treated with ivermectin (n=173) had better outcomes compared to those without ivermectin (p=0.03), even in patients with severe pulmonary involvement [12].

There is however some cause for concern amongst the experts, concerning drug safety due to ivermectin’s cytotoxicity. For this concern, liposomal drug delivery systems have been engineered which result in lesser cytotoxicity [13]. Ivermectin has been trialed at several doses ranging from low dose (150 μg/kg) to very high doses (120 mg/day) [14] and has been documented to be safe at higher doses in healthy volunteers. Although Schmith et al. [15] proposed that ivermectin is unlikely to reach IC50 in the lungs at standard approved dosing, our trial shows that the 12 mg dose not only delivers promising results but also without the development of any immediate side effects. However, to completely comprehend the safety profile further trials are needed.

There were however certain limitations and shortcomings of the study. The duration and severity of individual symptoms and time of resolution of these symptoms were not studied. Most of the patients were lost to follow up after the trial period concluded and very few could be traced back to assess for any potential adverse reaction that may have occurred due to treatment with ivermectin, hence prolonged safety of drug could not be established. However, no side effects were noted during the trial period and ivermectin was well tolerated. Majorly our participants were males and the results may not be generalizable to both genders.

Although our research gives an indicator to a very strong potential candidate for the treatment of COVID-19, all possible confounders can never be reliably ruled out. However, as the cases of COVID-19 surge, we have a reason to believe that even this initial information could be of importance for clinicians and researchers. This drug has an established therapeutic benefit and should be considered for testing in other setups. In continuation of this trial, enrollment needs to be extended to patients with severe symptoms of COVID-19, to reduce the need for treatment escalation.

## Conclusions

In the intervention arm, early viral clearance was observed in patients without experiencing any side effects. These are of importance because high viral load and prolonged viremia can potentially trigger the immune dysregulation phase leading to more severe disease, and the requirement of treatment escalation.

## Data Availability

The data can be made available from authors

## Declarations

### Ethical approval and consent to participate

the approval was sought from Combined Military Hospital Lahore Ethics Review Board (ERC # 169/2020) and the trial was registered before initiation (Trial registration # NCT04392713).

### Consent for publication

not applicable

## Acknowledgments

The authors would like to thank Dr Manahil Chaudhry for her technical support and helping in manuscript preparation.

## References

1. Campbell WC, Benz GW: Ivermectin: a review of efficacy and safety. J Vet PharmacolTher. 1984, 7:1–16. 10.1111/j.1365-2885.1984.tb00872.x

2. Yang SNY, Atkinson SC, Wang C, Lee A, Bogoyevitch MA, Borg NA, Jans DA: The broad spectrum antiviral ivermectin targets the host nuclear transport importin α/β1 heterodimer. Antiviral Res. 2020, 2:104760. 10.1016/j.antiviral.2020.104760

3. Chen, I.-S. and Kubo, Y. (2018: Ivermectin and its target molecules: shared and unique modulation mechanisms of ion channels and receptors by ivermectin. J Physiol. 596:1833–1845. 10.1113/JP275236

4. Banerjee K, Nandy M, Dalai CK, Ahmed SN: The Battle against COVID 19 Pandemic: What we Need to Know Before we “Test Fire” Ivermectin. Drug Res (Stuttg. 2020, 70:337–340. 10.1055/a-1185-8913

5. Heidary F, Gharebaghi R: Ivermectin: a systematic review from antiviral effects to COVID-19 complementary regimen. J Antibiot (Tokyo. 2020, 73:593–602. 10.1038/s41429-020-0336-z

6. Caly L, Druce JD, Catton MG, Jans DA, Wagstaff KM: The FDA-approved drug ivermectin inhibits the replication of SARS-CoV-2 in vitro. Antiviral Res. 2020 Jun, 178:104787. 10.1016/j.antiviral.2020.104787

7. Ye Q, Wang B, Mao J: The pathogenesis and treatment of the ‘Cytokine Storm’ in COVID-19. J Infect. 2020, 80:607–613. 10.1016/j.jinf.2020.03.037

8. Omrani AS, Saad MM, Baig K, et al.: Ribavirin and interferon alfa-2a for severe Middle East respiratory syndrome coronavirus infection: a retrospective cohort study [published correction appears in. Lancet Infect Dis. 2015 Jan, 13:1090– 1095. 10.1016/S1473-3099(14)70920-X

9. D.A. Jans, K.M: Wagstaff, The broad spectrum host-directed agent ivermectinas an antiviral for SARS-CoV-2?. Biochemical and Biophysical Research Communications. 10.1016/j.bbrc.2020.10.042

10. Coronavirus Disease 2019 (COVID- 19): A Perspective from China Zi Yue Zu, Meng Di Jiang, Peng Peng Xu, Wen Chen, Qian Qian Ni, Guang Ming Lu, and Long Jiang Zhang: Radiology. 2020, 296:15–25. 10.1148/radiol.2020200490

11. Bray M, Rayner C, Noël F, Jans D, Wagstaff K. Ivermectin and COVID- 19: A report in Antiviral Research, widespread interest, an FDA warning, two letters to the editor and the authors’ responses. Antiviral Res. 2020, 178:104805. 10.1016/j.antiviral.2020.104805

12. Rajter JC, Sherman M, Fatteh N, et al.: ICON (Ivermectin in COvid Nineteen) study: Use of Ivermectin is Associated with Lower Mortality in Hospitalized Patients with COVID19. medRxiv. 2020. DOI, 10.1101/2020.06.06.20124461

13. Sharun K, Dhama K, Patel SK, et al.: Ivermectin, a new candidate therapeutic against SARS-CoV-2/COVID-19. Ann Clin Microbiol Antimicrob. 2020;19(1, 23– 2020. 10.1186/s12941-020-00368-w

14. Guzzo CA, Furtek CI, Porras AG, et al.: Safety, tolerability, and pharmacokinetics of escalating high doses of ivermectin in healthy adult subjects. J Clin Pharmacol. 42:1122–1133. 10.1177/009127002401382731

15. Schmith, V.D., Zhou, J.(. and Lohmer, L.R. (2020: The Approved Dose of Ivermectin Alone is not the Ideal Dose for the Treatment of COVID-19. Clin. Pharmacol. Ther. 108:762–765.

